# Heterogeneity in paediatric diabetes clinical presentation and its link to disease severity

**DOI:** 10.1101/2025.11.10.25339758

**Authors:** Miguel A Juárez Garzón, Shania I Muganga, Divya Sri Priyanka Tallapragada, Bente B Johansson, Janne Molnes, Torild Skrivarhaug, Valeriya Lyssenko, Miriam Udler, Stefan Johansson, Ksenia G Kuznetsova, Marc Vaudel, Pål R Njølstad

## Abstract

**Background:** Diabetes subtypes with different clinical profiles have been found and replicated in adults. However, clinical heterogeneity of paediatric-onset diabetes remains unexplored. Better capture of different aetiologies of the disease could prove a powerful tool towards precision medicine.

**Methods:** We performed data-driven clustering analysis in patients with newly diagnosed diabetes (n = 3,064) from the Norwegian Childhood Diabetes Registry. Patients were stratified by autoantibody status and clustered based on five clinical variables: Age at diagnosis, fasting glucose, HbA1c, fasting C-peptide Z-score, and BMI Z-score. We assessed inter-cluster differences regarding severity of disease, early treatment needs, polygenic scores (PS), and serum biomarkers.

**Findings:** We identified two clusters of autoantibody-positive and three clusters of autoantibody-negative patients: 1) Childhood severe autoimmune diabetes (CSAID; n = 1482, 48·37 %), characterized by childhood-onset diabetes, milder disease presentation, unaltered lipidic profiles, higher Type 1 diabetes PS, and autoantibody positivity; 2) Adolescence severe autoimmune diabetes (ASAID; n = 1252, 40·86 %) with adolescent-onset of diabetes, more severe disease presentation, higher diabetic ketoacidosis, lipidic profiles signaling towards prolonged metabolic disease, and autoantibody positivity; 3) Childhood severe insulin-deficient diabetes (CSIDD; n = 106, 3·46 %) and 4) Adolescence severe insulin-deficient diabetes (ASIDD; n = 144, 4·70 %) with similar characteristics to CSAID and ASAID, respectively, but no autoantibody positivity; and 5) Adolescence severe insulin-resistant diabetes (ASIRD; n = 80, 2·61 %), the oldest group, with the highest C-peptide, BMI Z-score, and Type 2 diabetes PS. CSAID and ASAID presented similarities to previously described endotypes for type 1 diabetes.

**Interpretation:** We grouped patients with paediatric diabetes into five subgroups with varying clinical severity, genetic risk, and metabolic profiles. Similarities between autoantibody-positive and -negative clusters underscore the importance of adopting a personalised, multivariate approach to diabetes management that extends beyond autoantibody status. We hypothesise that our clusters may be connected to previously described endotypes of type 1 diabetes, facilitating patient classification without the need for pancreatic biopsies. Further understanding of this concept could help define the mechanisms involved in disease initiation, time to diagnosis, and progression.

## Introduction

Diabetes is a heterogeneous disease in its clinical presentation and progression. Patients are ascribed to subtypes represented by distinct clinical characteristics and underlying causes. The main subtypes are type 1 diabetes (T1D), type 2 diabetes (T2D), gestational diabetes (GDM), and monogenic diabetes, with maturity-onset diabetes of the young (MODY) being the most common form.^1^ This poses a challenge for disease management as treatment response is patient-specific. As a result, there is a growing interest in precision medicine approaches to identify patient subtypes. In adults, Alhqvist et al^2^ and later related studies have implemented unsupervised clustering analysis to identify subtypes of newly onset adult diabetes.^3^

For children, different endotypes have recently been described in patients with type 1 diabetes, based on analysis of pancreatic biopsies.^4,5,6^ However, this approach requires complex variables derived from the analysis of the biopsies. As a consequence, paediatric diabetes remains underexplored.^7^ In the present study, we used the Norwegian Childhood Diabetes Registry (NCDR) to identify clusters of paediatric diabetes using routine clinical variables, and studied these subgroups clinically, genotypically, and metabolically.

## Methods

### Study Population

Data and samples used in this study were derived from the Norwegian Childhood Diabetes Registry (NCDR),^8^ a prospective nationwide medical registry that includes children and adolescents below 18 years with all forms of diabetes. We used a sample that consisted of 3,064 individuals with newly diagnosed diabetes between the ages of 1 and 18 years from 2006 to 2018. A list of all variables used, including their means and interquartile ranges (IQR), is provided in Supplementary Table 1.

### Measurements and procedures

Clinical measurements and laboratory values were available from NCDR, as reported from various medical units across Norway. Patient selection was based on data completeness for age at diagnosis, fasting glucose, HbA1c, fasting C-peptide, and BMI. Extreme outliers (>5 SD from the mean) were filtered out. Patients were classified as autoantibody-positive when at least one of four autoantibodies (GAD65A, IA-2A, ZnT8A, and Insulin) was detected; the relevance of autoantibodies against insulin is still debated, however, we decided to include it based on its use in other studies of type 1 diabetes and its inclusion in the ADA Diagnosis and Classification criteria.^1^ Individuals with a confirmed genetic diagnosis harboring pathogenic or likely pathogenic variants related to monogenic diabetes were excluded from the clustering analysis.

Age- and sex-Z-scores were computed for BMI using reference values from Júlíusson et al,^9^ and reference values from various pediatric population studies for C-Peptide.^10,11,12^ Z-score computation was performed as described in the WHO Child Growth Standards: Methods and development book.^13^

### Genotyping and polygenic scores

Genotyping of 2,957 NCDR participants was performed on frozen DNA samples prepared from blood. Genome-wide genotyping of 741,178 markers was performed using the Illumina Infinum Global Screening Array 24 v2.0 by the Human Genomics Facility at Erasmus MC (Rotterdam, the Netherlands). Polygenic scores for Type 1 diabetes (PGS000833) and Type 2 diabetes (PGS000832) were computed from the genotyping data. Type 1 diabetes polygenic score was computed as a whole as well as split into its HLA and non-HLA components.

### Nuclear magnetic resonance (NMR) metabolomics

Metabolomic biomarkers from serum samples of 248 individuals were quantified using Nightingale Health NMR Core Metabolomics (Nightingale Health Plc, Helsinki, Finland). This assay provides measurements of routine lipids, apolipoproteins, lipoprotein subclass profiling, fatty acid composition, and various low-molecular metabolites, including amino acids, ketone bodies, and glycolysis-related metabolites.^14,15^

The raw data matrix with metabolite concentrations was filtered by removing all metabolites having more than 20% missing values across all samples. The remaining metabolites were imputed using the missForest R package version 1.5 .^16^ Imputed metabolomics were then log10-transformed and scaled using Pareto (mean-centered and divided by the square root of the standard deviation). Imputed and scaled metabolomics were used for further analysis.

### Clustering models implementation and validation

The clustering strategy was defined by testing K-means clustering and different forms of agglomerative hierarchical clustering using Euclidean distances. Males and females were initially clustered separately, but showed no differences and were pooled into a single analysis. Antibody-positive and antibody-negative patients were clustered separately.

The different clustering algorithms were tested with a range of clusters from two to ten. We computed validation metrics (Connectivity, Dunn, and Silhouette) to rank the different algorithms and the number of clusters. To validate clusters, we computed the Jaccard stability index and accepted only those solutions with a Jaccard index above 0·75 in all clusters. The best algorithm was selected as that with the best metrics and with a Jaccard Index above 0·75 for all clusters (Supplementary Table 2 & 3). The NcbClust package was then used to verify the best number of clusters for each case. After this step, k-means clustering with two and three clusters for antibody-positive and antibody-negative patients, respectively, was retained. Furthermore, we verified that all variables selected as input in the clustering analysis were significantly different between at least two clusters.

### Statistical analysis

Continuous variables were checked for normal distribution via the Shapiro-Wilk Test. Normally distributed variables were analyzed using Student’s t-test when comparing two clusters or ANOVA followed by Fisher’s LSD post hoc test when analyzing three clusters. Non-normally distributed variables were analysed using Kruskal-Wallis for both two and three clusters, and Dunn post hoc test was used when comparing three clusters. Categorical variable significance was assessed using χ2. Metabolite significance was assessed using logistic regression between cluster pairs. The metabolite significance threshold was corrected for multiplicity (α = 0·05/139) using the Nyholt method.^17,18^All statistical analyses were performed using R version 4.4.2. For all tests, the two-sided significance level was set to 0·05 unless otherwise stated.

## Results

### Unsupervised clustering identifies five subtypes of paediatric diabetes

We first tested different clustering algorithms on 3,064 children from NCDR. All individuals had complete data for the clinical variables: age at diagnosis, fasting glucose, HbA1c, fasting C-peptide Z-score, and BMI Z-score. Antibody-positive and antibody-negative patients were clustered separately. The k-means algorithm performed best in both cases. The optimal number of clusters was k = 2 and k = 3 for patients classified as antibody-positive and -negative, respectively (Supplementary Figure 1).

Based on this, we defined five main clusters: Childhood severe autoimmune diabetes (CSAID), characterized by onset during childhood, autoantibody positivity, and reduced BMI and C-peptide levels, comprising 1482 (48·37 %) patients; adolescent severe autoimmune diabetes (ASAID), showing autoantibody positivity, high glucose and HbA1c values, low BMI and C-peptide z-scores, and, including 1252 (40·86 %) patients; childhood severe insulin-deficient diabetes (CSIDD), characterized by onset during childhood, no detectable autoantibodies, and reduced BMI and C-peptide levels, similar to CSAID, encompassing 106 (3·46 %); adolescent severe insulin-deficient diabetes (ASIDD), with diagnosis during adolescence, no detectable autoantibodies, high glucose and HbA1c values, and low BMI and C-peptide z-scores, consisting of 144 (4·70 %); and adolescent severe insulin-resistant diabetes (ASIRD), featuring the latest age at diagnosis, highest BMI Z-score and C-peptide Z-score and the lowest glucose and HbA1c, comprising 80 (2·61 %) patients (Figure 1; Supplementary Table 4). The couples of clusters CSAID-CSIDD and ASAID-ASIDD presented similar distributions for all variables.

**Figure 1.**
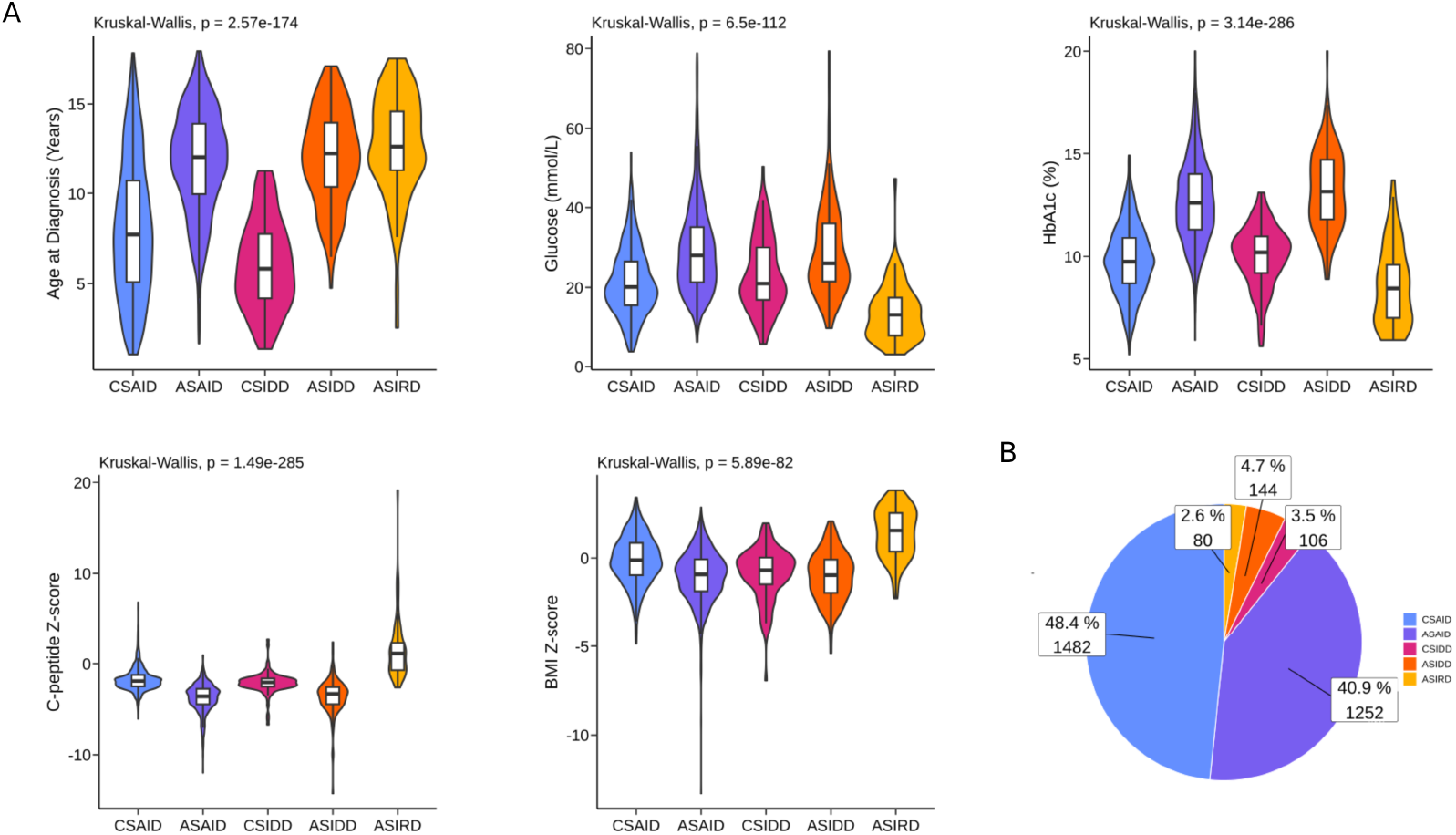
Pediatric diabetes clusters in the NCDR cohort. (A) Distribution of clustering variables: Age at diagnosis, fasting glucose, HbA1c, C-peptide Z-score, and BMI Z-score in clusters of the NCDR cohort. (B) Participant percentage and total number per cluster in NCDR. CSAID = childhood severe autoimmune diabetes. ASAID = adolescence severe autoimmune diabetes. CSIDD = childhood severe insulin-deficient diabetes. ASIDD = adolescence severe insulin-deficient diabetes. ASIRD = adolescence severe insulin-resistant diabetes. NCDR = Norwegian Childhood Diabetes Registry.

### Adolescence diabetes shows increased diabetic ketoacidosis and early insulin needs

Inter-cluster differences were further studied using clinical variables collected during admission to the clinics and trajectories recorded during stay in the hospital. Regarding acid-base blood balance, ASAID and ASIDD showed significantly lower levels of both pH and bicarbonate when compared to CSAID, CSIDD, and ASIRD. Despite CSAID, CSIDD, and ASIRD pH levels being similar, ASIRD bicarbonate levels were significantly increased (Figure 2A). In addition, ASAID and ASIDD showed higher blood and urine ketone levels compared to CSAID, CSIDD, and ASIRD (Figure 2B). Collectively, these results demonstrate a more severe disease presentation in ASAID and ASIDD.

**Figure 2.**
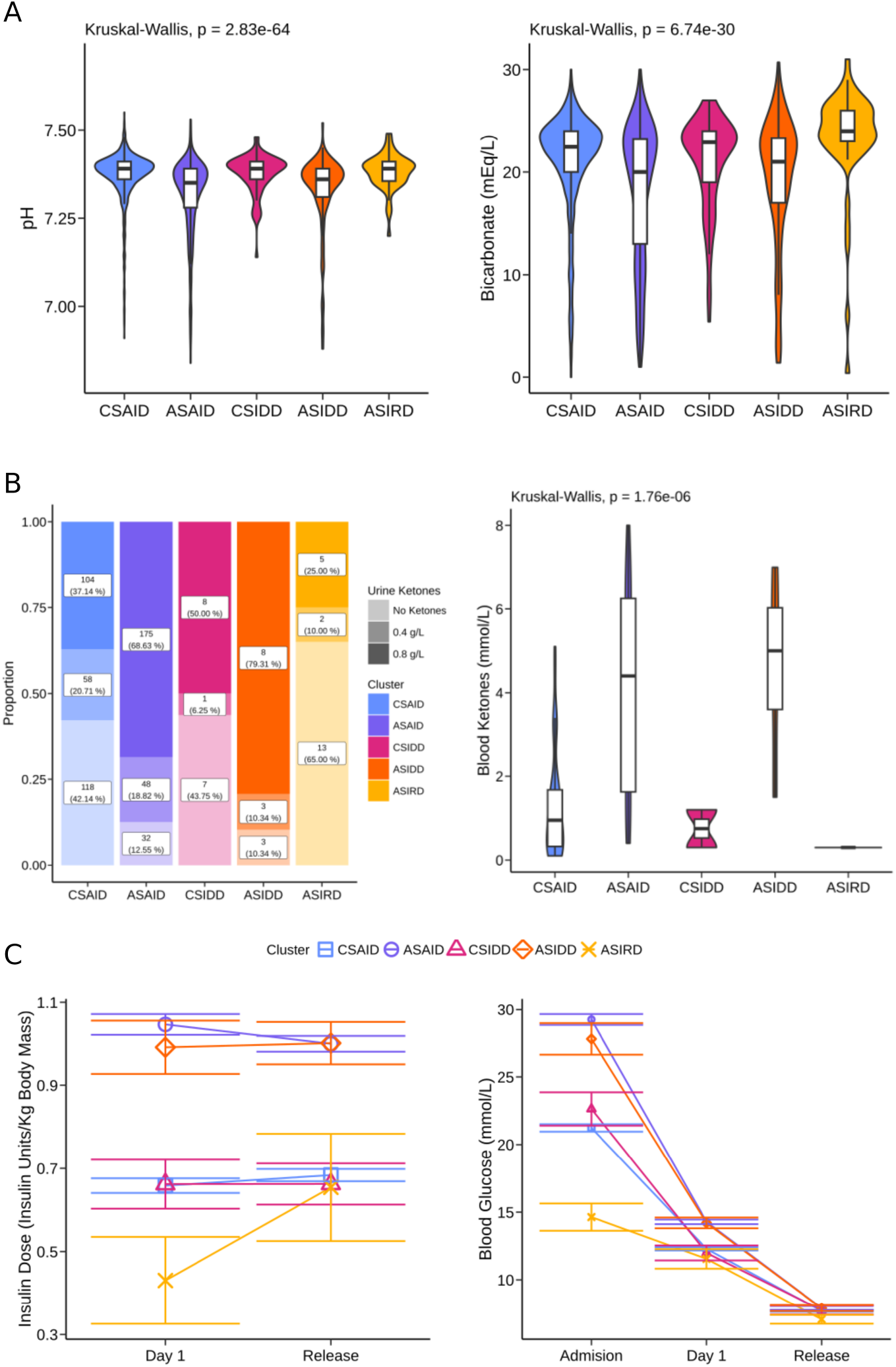
Clinical variables at diagnosis and progression during hospital admission. (A) Blood acid-base balance measured by pH and bicarbonate in the NCDR cohort. (B) Urine and blood ketone bodies in the NCDR cohort. (C) Fasting glucose and insulin dosage progression during hospital admission. CSAID = childhood severe autoimmune diabetes. ASAID = adolescence severe autoimmune diabetes. CSIDD = childhood severe insulin-deficient diabetes. ASIDD = adolescence severe insulin-deficient diabetes. ASIRD = adolescence severe insulin-resistant diabetes. NCDR = Norwegian Childhood Diabetes Registry.

Furthermore, CSAID, CSIDD, and ASIRD displayed significantly lower insulin needs both one day after admission and at release, as well as lower glucose levels compared to ASAID and ASIDD. ASIRD presented the lowest glucose levels of all clusters. While differences in insulin needs remained relatively unchanged during the hospital stay, glucose levels normalized and by the time of release, no significant difference could be observed between clusters (Figure 2C). As observed in the cluster variable analysis, the CSAID-CSIDD and ASAID-ASIDD clusters were paired, showing similar distributions for all variables studied.

For autoantibody-positive patients, no significant differences were observed between CSAID and ASAID regarding the number of autoantibodies detected at diagnosis when analysing patients with complete data for all autoantibodies. However, the proportion of anti-insulin autoantibodies was slightly elevated in CSAID, while the proportion of anti-GAD autoantibodies was increased in ASAID (Supplementary Figure 2).

### Difference in genetic risk between childhood and adolescence diabetes clusters is driven by HLA SNPs

Genetic differences between clusters were examined using T1D and T2D polygenic scores (PS) computed from SNP-based genotyping. This analysis was performed on a subset of 2,957 individuals. CSAID, ASAID, CSIDD, and ASIDD showed higher T1D PS and low T2D PS, while ASIRD showed the lowest T1D PS and highest T2D PS (Figure 3A-B). Furthermore, CSAID and CSIDD showed a slightly higher T1D PS than ASIDD, with this difference being driven by the HLA component of the PS (Figure 3C). As previously observed with phenotypic variables, the pairs of clusters CSAID-CSIDD and ASAID-ASIDD presented similar distributions for both T1D and T2D PSs.

**Figure 3.**
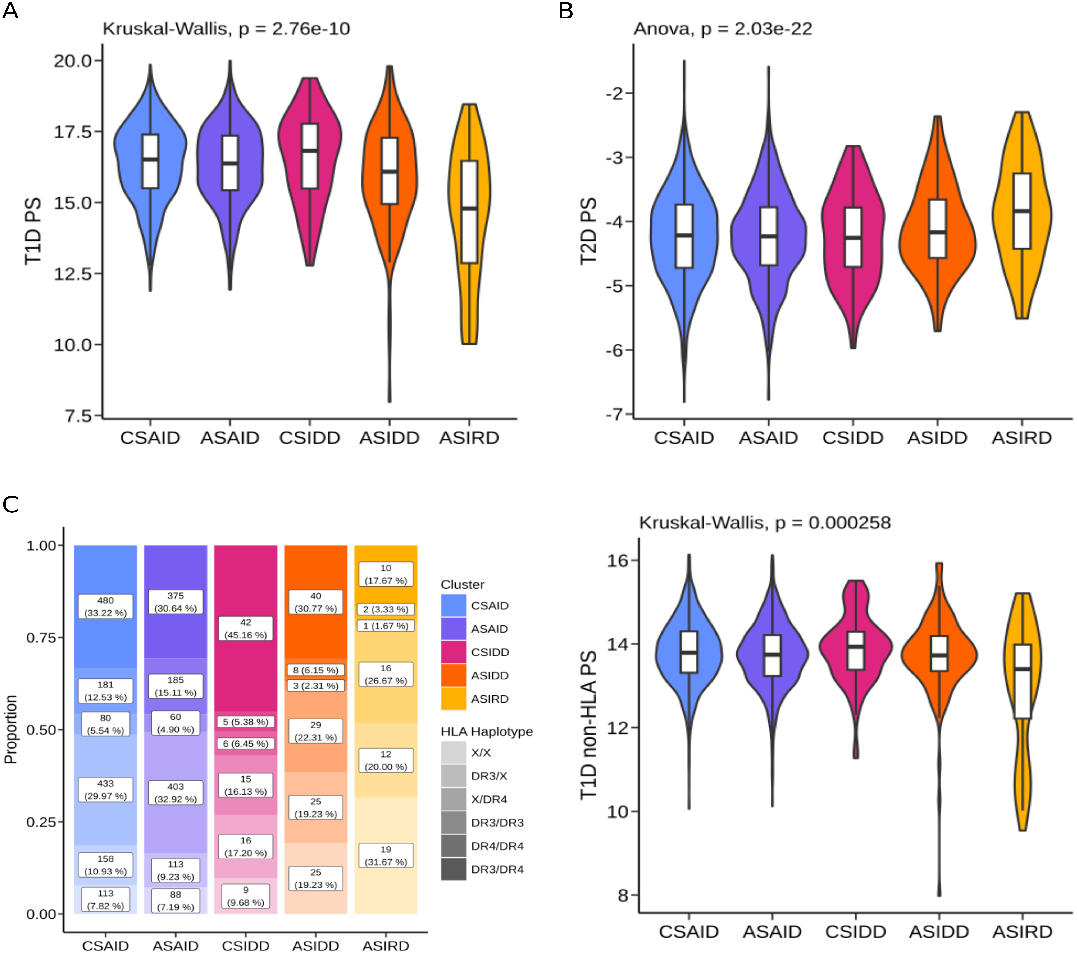
Polygenic risk scores by cluster in the NCDR cohort. (A) Type 1 diabetes polygenic score (B) Type 2 diabetes polygenic score (C) Type 1 HLA and Type 1 non-HLA polygenic score distribution in NCDR clusters. Kruskal-Wallis, Anova or Chi-Square P-values displayed. CSAID = childhood severe autoimmune diabetes. ASAID = adolescence severe autoimmune diabetes. CSIDD = childhood severe insulin-deficient diabetes. ASIDD = adolescence severe insulin-deficient diabetes. ASIRD = adolescence severe insulin-resistant diabetes. NCDR = Norwegian Childhood Diabetes Registry.

### Serum metabolic variation in paediatric diabetes

Differences in metabolic profiles between clusters were analyzed using NMR measurements of 182 metabolites in samples from 248 patients. Multiplicity correction based on the Nyholt method (*α* = 0·05/139) was applied. Significant differences were observed only when comparing CSAID and CSIDD against ASAID. In both cases, ASAID showed increased levels of triglycerides (Supplementary Figure 3).

Based on the lack of statistical power and the similarities in phenotypic, genetic, and metabolic profiles between the CSAID-CSIDD and ASAID-ASIDD couples, we decided to pool these clusters to increase the statistical power of our metabolomic analysis. The pooled metabolomic analysis revealed significant differences between CSAID-CSIDD and ASAID-ASIDD, with individuals in the ASAID-ASIDD cluster exhibiting increased levels of triglyceride molecules of all sizes. Creatinine was significantly increased in the ASAID-ASIDD cluster, while albumin was higher in the CSAID-CSIDD cluster (Figure 4). No significant differences were observed between ASIRD and CSAID-CSIDD or ASAID-ASIDD, which may be due to insufficient statistical power resulting from the low number of individuals in the ASIRD cluster (Supplementary Figure 4). Overall, these results show changes related to poor glycemic control and metabolic stress, and early signs of renal dysfunction in the ASAID-ASIDD subgroup.

**Figure 4.**
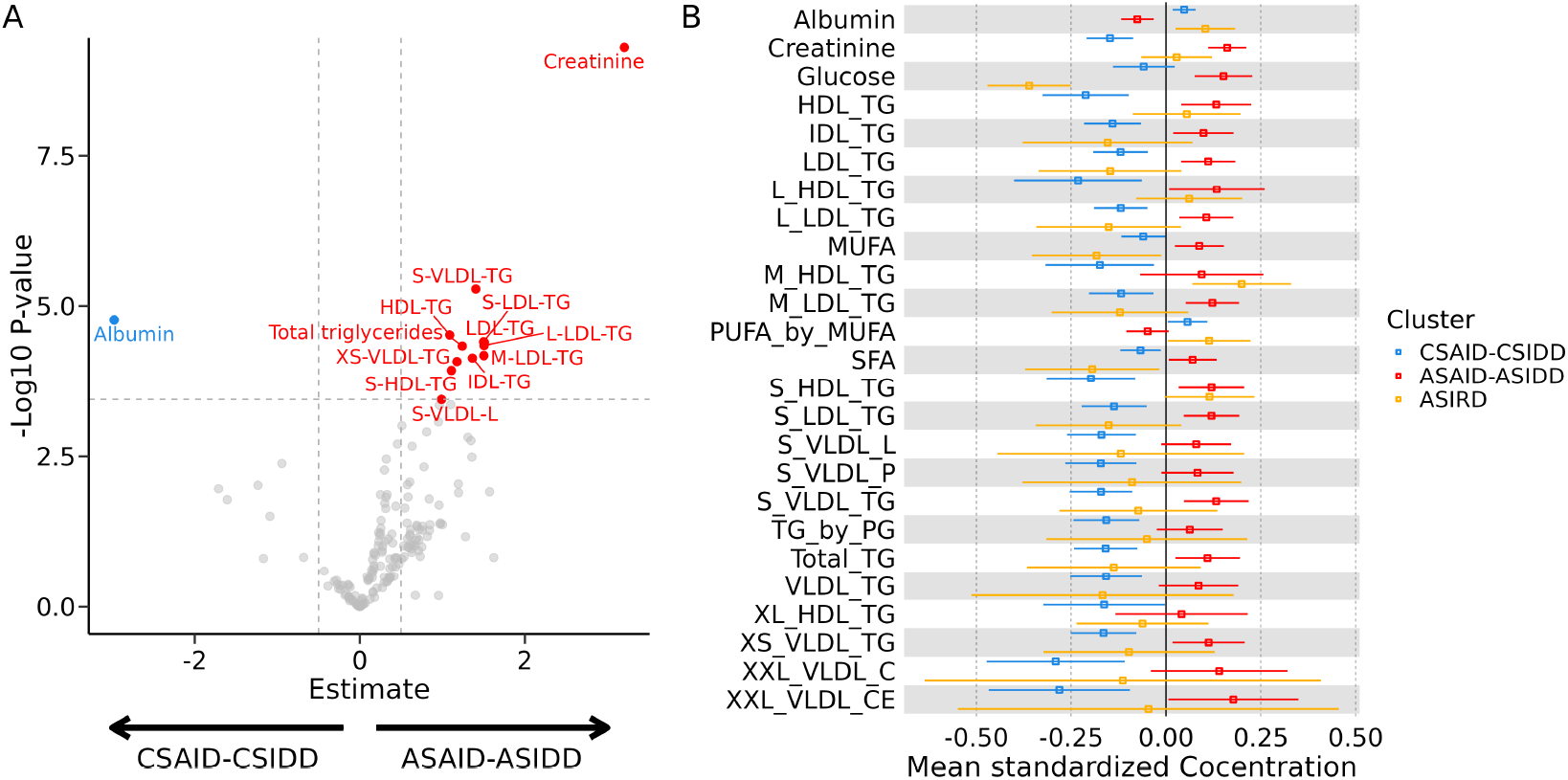
Metabolomic comparison of NCDR cohort clusters. (A) Comparison between CSAID-CSIDD and ASAID-ASIDD metabolites. Forestplot of the top 25 metabolites ordered by p-value of Kruskal-Wallis or ANOVA analysis. CSAID = childhood severe autoimmune diabetes. ASAID = adolescence severe autoimmune diabetes. CSIDD = childhood severe insulin-deficient diabetes. ASIDD = adolescence severe insulin-deficient diabetes. ASIRD = adolescence severe insulin-resistant diabetes. NCDR = Norwegian Childhood Diabetes Registry.

### Projection of monogenic diabetes cases in childhood diabetes clusters

We tested how 24 patients, external to the clustering analysis, with genetic diagnosis for mutations in six different MODY genes (GCK-MODY, HNF1A-MODY, HNF1B-MODY, CEL-MODY, INS-MODY, and ABCC8-MODY) would project in our clustering. For this, we computed distances to cluster centroids and assigned each case to the closest cluster. All patients with GCK-MODY, HNF1A-MODY, and INS-MODY were assigned to ASIRD. Patients with HNF1B-MODY were assigned to ASIDD. Finally, patients diagnosed with ABCC8-MODY and one patient with CEL-MODY, all of whom were autoantibody positive, were distributed between ASAID and CSAID (Figure 5).

**Figure 5.**
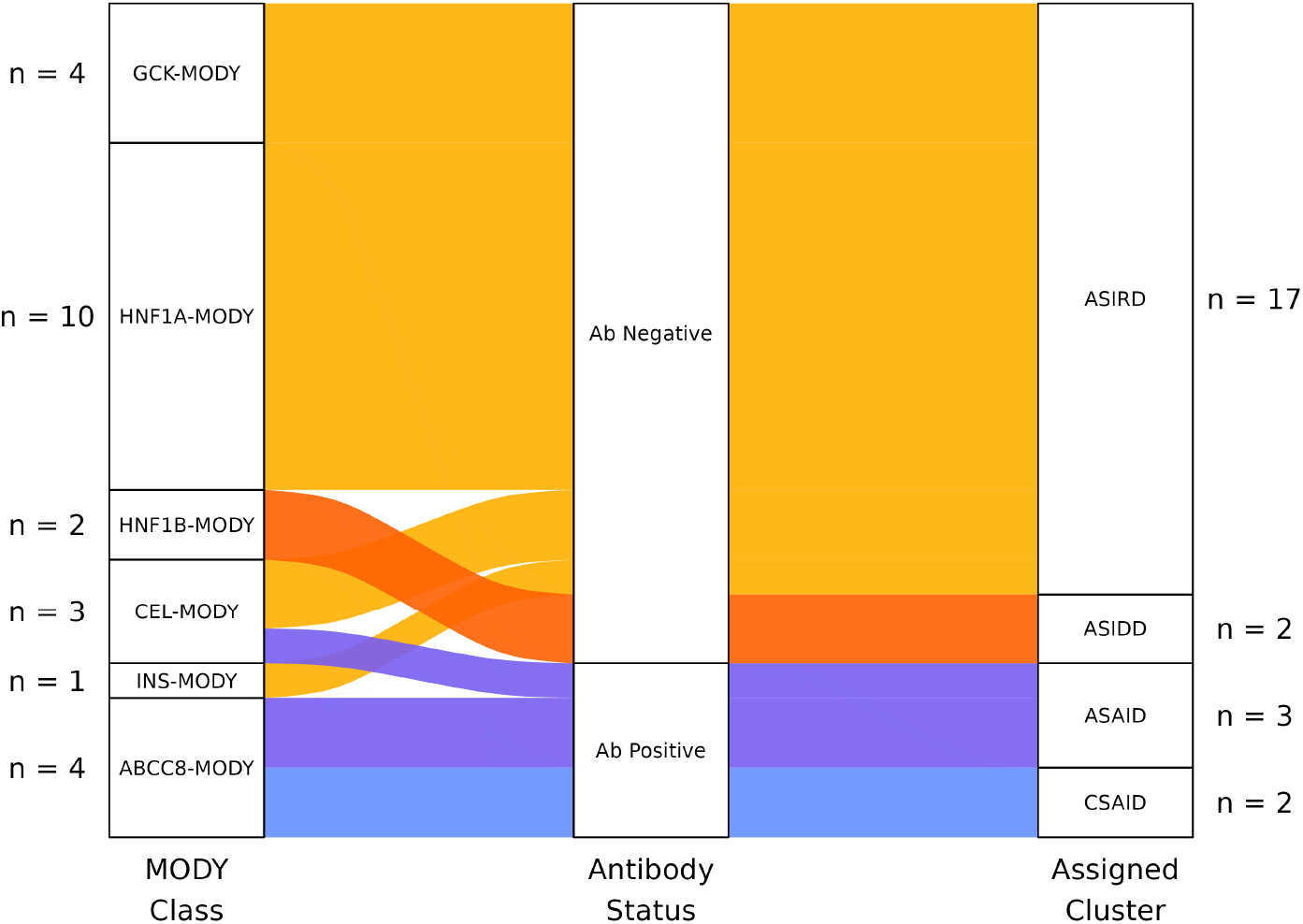
Projection of MODY cases in the NCDR clusters. Line color represents the assigned cluster of each case. MODY = maturity-onset diabetes of the young. CSAID = childhood severe autoimmune diabetes. ASAID = adolescence severe autoimmune diabetes. CSIDD = childhood severe insulin-deficient diabetes. ASIDD = adolescence severe insulin-deficient diabetes. ASIRD = adolescence severe insulin-resistant diabetes. NCDR = Norwegian Childhood Diabetes Registry.

## Discussion

This study aimed to investigate the heterogeneity of patients with pediatric diabetes using routine clinical variables. We identified a five-cluster subdivision. The ASIRD cluster was formed by 2·5% of the patients, characterized by increased BMI and C-peptide production, and the highest T2D PS. All phenotypic characteristics, PS, and the percentage of patients fit those already observed in youth-onset type 2 diabetes.^19^ The CSAID and ASAID clusters showed low BMI and C-peptide levels and high prevalence of autoantibodies, characteristic of type 1 diabetes, but they presented differences in their clinical presentation and age at diagnosis. CSIDD and ASIDD presented similar differences to those observed between CSAID and ASAID, but no autoantibodies could be detected. On the one hand, CSAID-CSIDD showed the highest T1D PS, which, as previously shown, can lead to earlier onset of the disease.^6,20^ On the other hand, ASAID-ASIDD increased blood glucose, HbA1c, and diabetic ketoacidosis (DKA) levels, point towards a prolonged exposure to severe hyperglycemia. In addition, ASAID-ASIDD metabolic profiles were shifted towards lipid usage, with triglyceride levels highly increased. This metabolic shift is commonly observed in patients with prolonged exposure to metabolic diseases, poor glycemic control, and DKA.^21,22,23^ All these results signal towards ASAID and ASIDD having a prolonged exposure to severe hyperglycemia, leading to extreme disease presentation, more severe outcomes, and metabolic consequences in the early stages post-diagnosis.

Similarities observed between CSAID-CSIDD and ASAID-ASIDD align with data reported in the ADA diagnosis guidelines, where 5-10% of patients with type 1 diabetes phenotype (low BMI and reduced C-peptide production) do not exhibit autoantibody positivity.^1^ Furthermore, Perchard et al^24^ already showed that in children, the antibody-positive and -negative individuals present no phenotypic differences, while adults have a different phenotypic presentation as shown in Ahlqvist et al.^2^ However, to our knowledge, this is the first time that paediatric patients with autoantibody negativity have been shown to display similarity with patients with antibody positivity, both phenotypically, genetically, and metabolically. However, the reasons for the lack of autoantibodies in these patients and the mechanisms behind disease initiation and progression remain unclear. As some studies suggest, immune processes independent of autoantibody presence or the reduced number of autoantibodies tested could be a reason for this.^25^ Several autoantigens have been observed to play a role in T1D immune response, either by CD4+ and CD8+ T cells attack, like IGRP or ChgA, or by generation of autoantibodies, like IAAP, Peripherin, or others.^26^ Further study of this subset of patients could help clarify the mechanisms behind this subgroup.

Furthermore, CSAID and ASAID align with the previously described endotypes for type 1 diabetes, type 1 diabetes endo-type 1 (T1DE1) and type 1 diabetes endotype 2 (T1DE2), as described by Leete et al.^5,6,27^CSAID and T1DE1 are both formed by patients diagnosed around six years of age and with a higher prevalence of anti-insulin autoantibodies. Furthermore, T1DE1 is characterized by a higher infiltration of immune cells in the pancreas as well as a higher presence of CD20+ lymphocytes and elevated proinsulin/C-peptide ratio. ASAID and T1DE2 are characterized by diagnosis after 13 years of age and a higher proportion of patients with anti-GAD autoantibodies. T1DE2 shows lower immune infiltration in the pancreas and normal proinsulin/C-peptide ratio. Based on these observations, we propose that patients in the CSAID cluster might present a stronger initial immune response, leading to earlier detection of symptoms and therefore shorter exposure to severe hyperglycemia. As a result, the initial phenotype and metabolic consequences would be milder for these patients. Conversely, patients in the ASAID cluster would have a milder immune response, leading to a progressive accumulation of the disease effects for a longer period, and a delay in the detection of symptoms, probably aggravated by the increased independence during adolescence. This would then lead to a more severe phenotype at diagnosis, less favorable initial prognosis, and metabolic changes. However, further analysis in prospective studies will be needed to demonstrate the correspondence of our clusters with previously described endotypes and the mechanisms of disease progression.

Regarding patients with a genetic diagnosis of MODY, we observed that almost all types of MODY mutations led to classification as part of the ASIRD cluster, except for patients with combined MODY and antibody positivity, or in the case of patients with an HNF1B-MODY diagnosis. Patients with HNF1B-MODY are usually treated with insulin in the first year following diagnosis, as a result of low pancreatic β-cell mass due to pancreatic malformation.^28,29,30^ These factors would explain their classification as ASIDD, as the reduced C-peptide levels and age at diagnosis match those of this cluster.

Based on the presented results, key strengths of our study include the use of a large, well-characterized, population-based cohort encompassing a wide spectrum of clinical presentations of paediatric diabetes, supported by an extensive set of phenotypic and biochemical variables. The simplicity of our clustering model and the use of data commonly available in the clinical setup as clustering variables make it easily replicable by clinicians. Furthermore, to our knowledge, this is the first clustering of patients with pediatric diabetes that looks at the entire population of patients with both antibody-positive and -negative diagnoses.

Further studies are needed to investigate whether our clusters correspond with the previously proposed endotypes for T1D and, therefore, with different aetiologies of the disease. Furthermore, differences between clusters after treatment initiation, and cluster transitions as well as their predictive ability for disease progression, need to be shown in future prospective studies, and the replicability of our study in populations of both European and other ancestries will need to be assessed in other cohorts. The stratification strategy could also be improved in the future by applying a multi-omic approach that combines different data modalities such as proteomic, genomic, or epigenomic markers.

In conclusion, our results suggest that a clustering strategy based on routine clinical variables can better capture the heterogeneity of paediatric diabetes than the classical autoantibody status classification, by grouping patients with similar clinical presentations. Furthermore, the combination of our clustering results with those previously described in adults lead to a continuous gradient of disease mechanisms and presentations from childhood until adulthood. This is an important step towards a precise stratification of paediatric diabetes, which we hope will be able to discern disease aetiologies, inform on disease progression, treatment options, and even open the door to new clinical trials targeting different groups of patients.

## Ethical declarations

Informed consent was obtained from all registry participants. The administrative board of the Norwegian Childhood Diabetes Registry approved the study protocol. The establishment of the Norwegian Childhood Diabetes Registry and initial data collection was based on a license from the Norwegian Data Protection Agency and approval from The Regional Committee for Medical Research Ethics. The Norwegian Childhood Diabetes Registry cohort is currently regulated by the Norwegian Health Registry Act. The study was approved by The Regional Committee for Medical Research Ethics (18836).

## Data availability

Data from the Norwegian Childhood Diabetes Registry can be made available to researchers, provided approval from the Regional Committees for Medical and Health Research Ethics (REC), compliance with the EU General Data Protection Regulation (GDPR) and approval from the data owners. The consent given by the participants does not open for storage of data on an individual level in repositories or journals. Researchers who want access to data sets for replication should apply to the registry. Access to data sets requires approval from The Regional Committee for Medical and Health Research Ethics in Norway and an agreement with the Norwegian Childhood Diabetes Registry (www.oslodiabetes.no/childhood).

## Code Availability

The code used to obtain the results presented in this manuscript is available athttps://github.com/ma-juarez/Pediatric_Diabetes_Clustering

## Acknowledgment

We thank all patients, personnel, and the Norwegian Childhood Diabetes Registry for the support and willingness to participate. M.V. was supported by the Research Council of Norway (#301178), the European Research Council (#101171420), and the University of Bergen. We also thank Liv Aasmul, Monika Ringdal and Louise Grevle for their technical and administrative support. All analyses were performed using digital labs in HUNT Cloud at the Norwegian University of Science and Technology, Trondheim, Norway. We are grateful for outstanding support from the HUNT Cloud community.

## Author contributions

MAJG, SIM, DSPT performed data analysis and interpretation. BBJ, JM, TS, VL, and MH helped with data analysis, provided expertise for interpretation of the results and writing of the manuscript. SJ, KK, MV, and PRN supervised analyses and interpretation. TS, BBJ, JM, SJ, KK, MV, and PRN led data acquisition and consolidation. MAJG, MV and PRN wrote the manuscript with input from all authors. All authors contributed to the manuscript edition and approved the final version of the manuscript. KK, MV, and PRN conceived the study and supervised the work.

