## Supplementary Figures for "Heterogeneity in paediatric diabetes clinical presentation and its link to disease severity"

#### Supplementary Figure Legends

**Supplementray Figure 1: Assessment of optimal cluster number in NCDR using clustering variables: Age at diagnosis, fasting glucose, HbA1c, C-peptide Z-score, and BMI Z-score** (A) Optimal number of clusters for autoantibody-positive patients. NbClust package output for the optimal number of clusters and Jaccard index. k=2 was the optimal cluster number. (B) Optimal number of clusters for autoantibody-negative patients. NbClust package output for the optimal number of clusters and Jaccard index. k=3 was the optimal cluster number.

**Supplementary Figure 2: Autoantibody distribution in CSAID and ASAID clusters for the NCDR cohort.** (A) Total number of autoantibodies detected in CSAID and ASAID clusters. (B) Proportion of positive and negative cases for Insulin, GAD, IA2A and ZnT8A autoantibodies. CSAID = childhood severe autoimmune diabetes. ASAID = adolescence severe autoimmune diabetes. NCDR = Norwegian Childhood Diabetes Registry.

**Supplementary Figure 3: Metabolomic comparison of NCDR cohort clusters.** (A) Comparison between CSAID and ASAID metabolites. (B) Comparison between CSAID and CSIDD metabolites. (C) Comparison between CSAID and ASIDD metabolites. (D) Comparison between CSAID and ASIRD metabolites. (E) Comparison between ASAID and CSIDD metabolites. (F) Comparison between ASAID and ASIDD metabolites. (G) Comparison between ASAID and ASIRD metabolites. (H) Comparison between CSIDD and ASIDD metabolites. (I) Comparison between CSIDD and ASIRD metabolites. (J) Comparison between ASIDD and ASIRD metabolites. CSAID = childhood severe autoimmune diabetes. ASAID = adolescence severe autoimmune diabetes. CSIDD = childhood severe insulin-deficient diabetes. ASIDD = adolescence severe insulin-deficient diabetes. ASIRD = adolescence severe insulin-resistant diabetes. NCDR = Norwegian Childhood Diabetes Registry.

**Supplementary Figure 4: Metabolomic comparison of merged NCDR cohort clusters.** (A) Comparison between CSAID-CSIDD and ASAID-ASIDD metabolites. (B) Comparison between CSAID-CSIDD and ASIRD metabolites. (C) Comparison between ASAID-ASIDD and ASIRD metabolites. CSAID = childhood severe autoimmune diabetes. ASAID = adolescence severe autoimmune diabetes. CSIDD = childhood severe insulin-deficient diabetes. ASIDD = adolescence severe insulin-deficient diabetes. ASIRD = adolescence severe insulin-resistant diabetes. NCDR = Norwegian Childhood Diabetes Registry.

**Supplementary Figure 1**

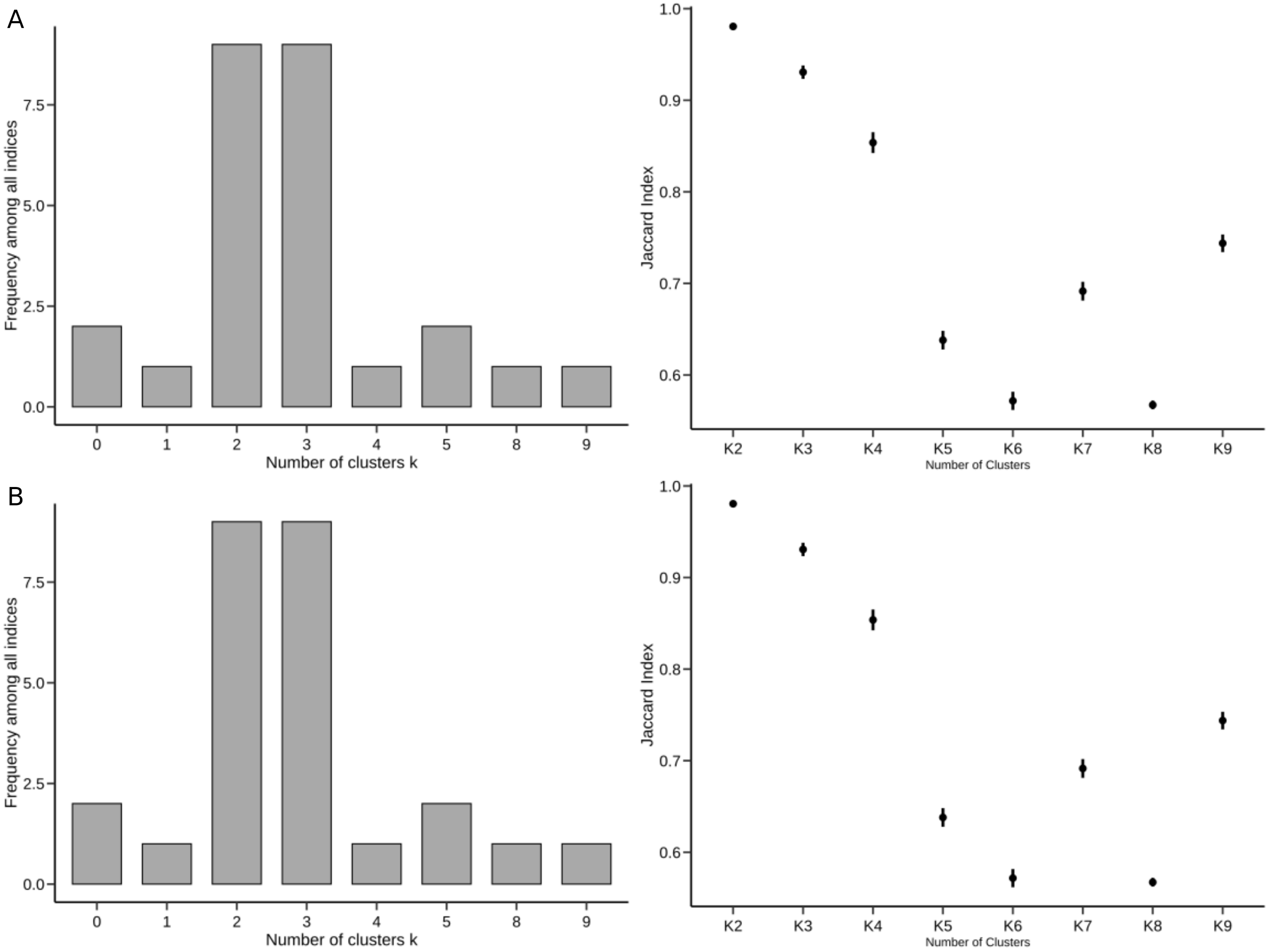

#### Supplementary Figure 2

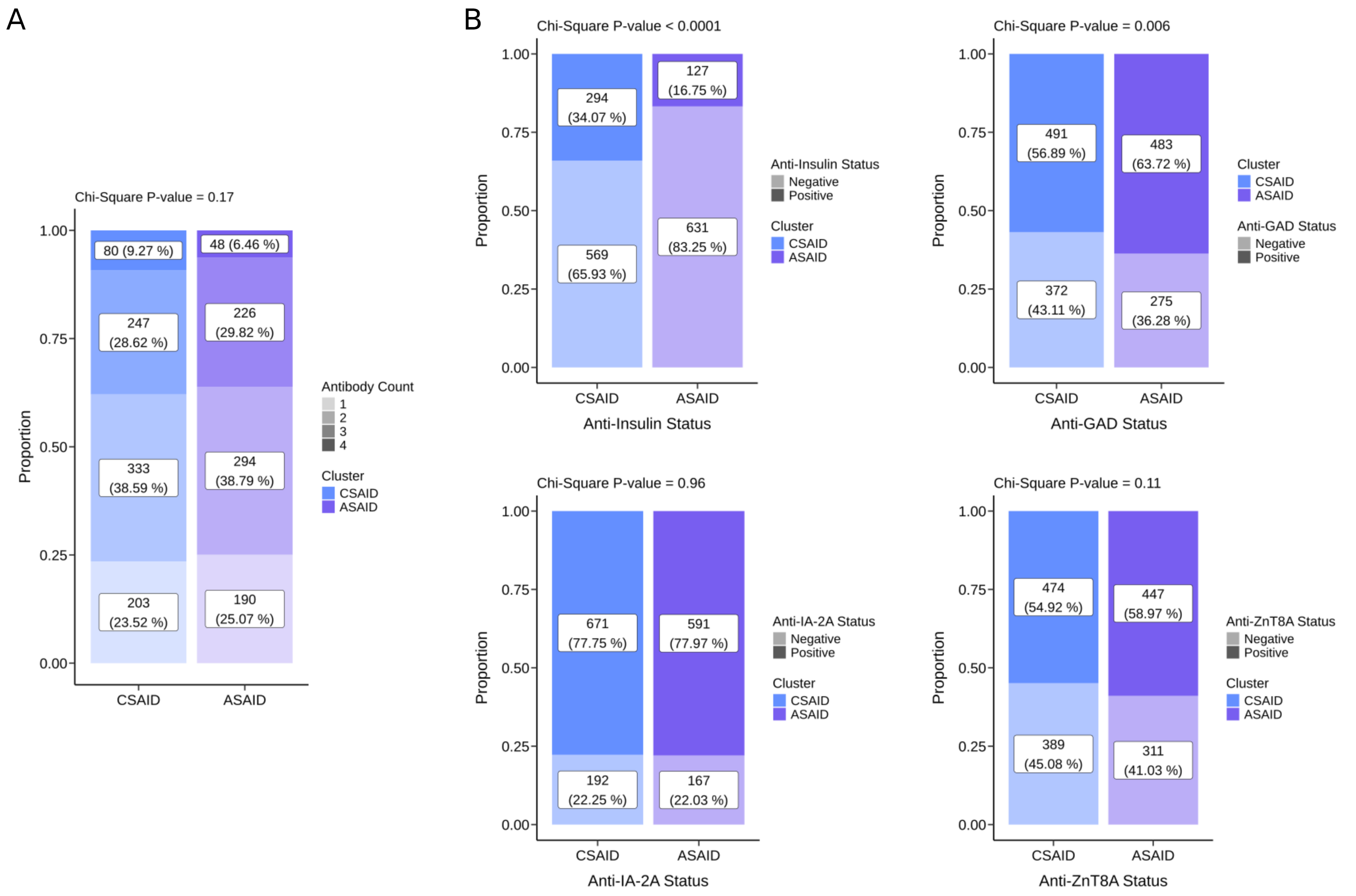

### Supplementary Figure 3

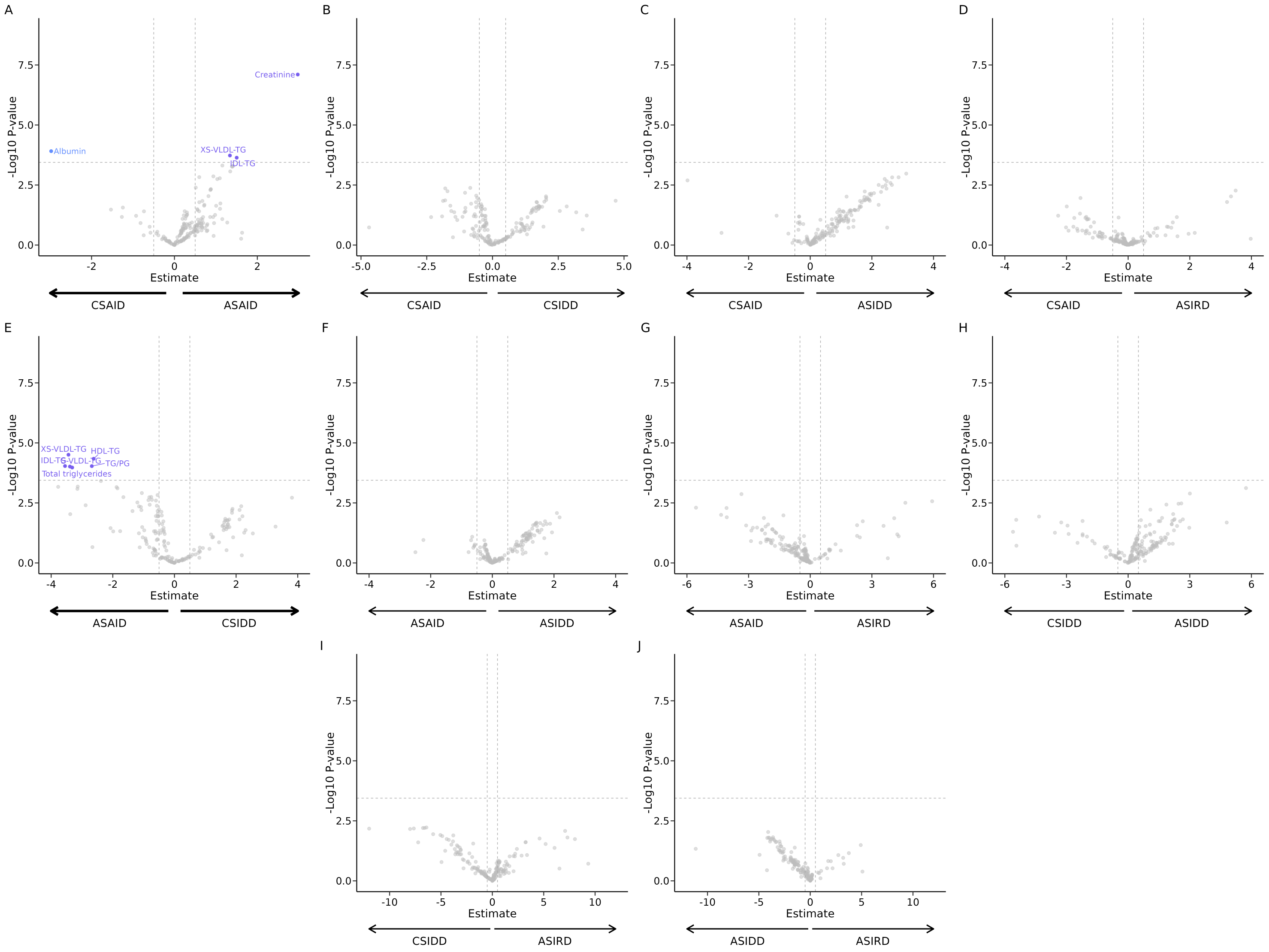

**Supplementary Figure 4**

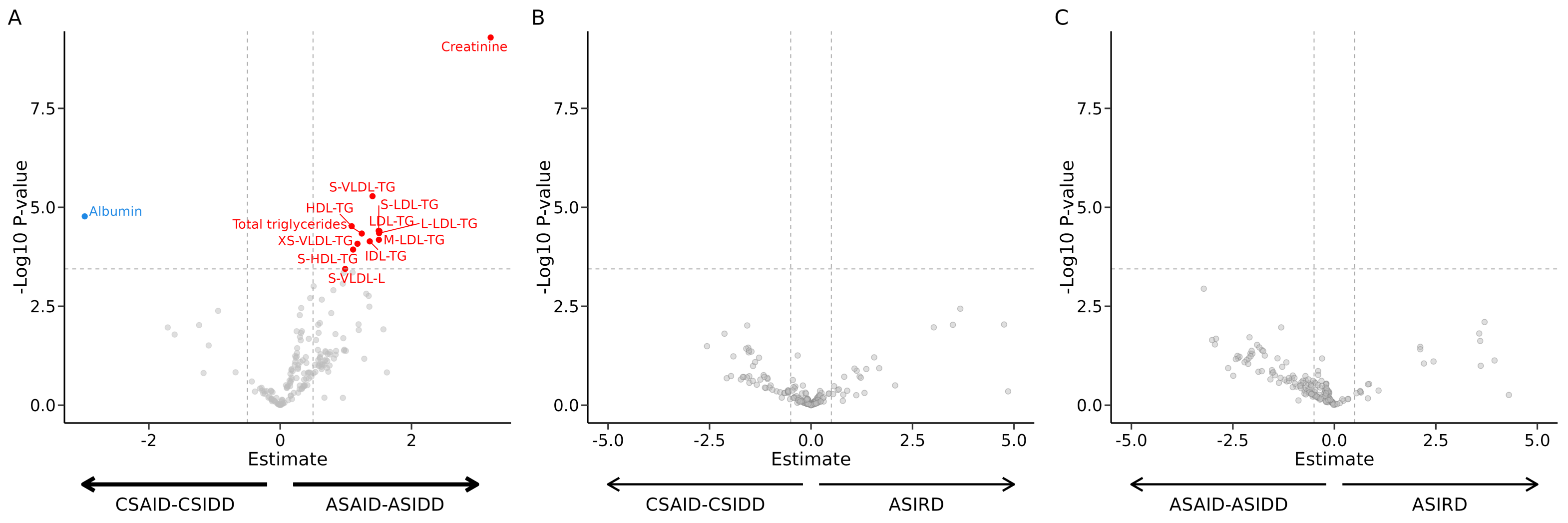
